# Prevalence of Hypertension among Indian Adults Based on Global Standards: Evidence from a nationally representative survey(NFHS-5)

**DOI:** 10.1101/2025.09.17.25335963

**Authors:** Harshvardhan Singh, Rishab Kumar Rana, Shailja Sharma, Sonu Goel

## Abstract

**Introduction:** Hypertension (HTN) is defined using varying systolic and diastolic thresholds across global guidelines and surveys. In India, the National Family Health Survey 5(NFHS 5) reports HTN among individuals >15 years, classifying it as mild, moderate, or severe, whereas international frameworks such as JNC 7, AHA, and ISH apply different cut offs for those >18 years. Such inconsistencies complicate clinical decision making, hinder policy formulation, and limit research comparability. This study aimed to assess the variability in HTN prevalence when applying different classification criteria and to underscore the need for a standardized framework.

**Methods:** We analyzed secondary data from NFHS 5, restricting the sample to men (18 to 54 years) and women (18 to 49 years). Analyses were performed using SPSS 29.0. Prevalence was reported with 95% confidence intervals (CI), and multivariate logistic regression estimated adjusted odds ratios (AORs). Pre hypertensives (JNC 7), elevated BP (AHA), and high normal BP (ISH) categories were excluded.

**Results:** NFHS 5 reported HTN prevalence of 21.3% in women (15 to 49 years) and 24% in men (15 to 54 years). Applying JNC/ISH cut offs to the same cohort yielded lower prevalence: 12.35% (95% CI: 12.27, 12.43) in women and 21.78% (95% CI: 21.49, 22.07) in men. By contrast, AHA criteria produced substantially higher estimates: 45.53% (95% CI: 45.41, 45.65) in women and 62.55% (95% CI: 62.22, 62.28) in men. Multivariate analysis showed significantly higher odds of HTN among individuals with BMI >25, lowest education, highest wealth quintile, alcohol use, smoking, and diabetes.

**Conclusions:** HTN prevalence varies markedly depending on the classification applied, revealing inconsistencies across NFHS, JNC 7, AHA, and ISH guidelines. This variability risks fragmented policy, inequitable treatment decisions, and reduced comparability of research. Establishing a uniform, globally accepted HTN classification framework is essential to strengthen surveillance, harmonize clinical practice, and accelerate progress toward reducing the global hypertension burden by 2030 (SDG).

## Introduction

Hypertension, commonly referred to as elevated blood pressure, is identified on the basis of defined systolic and diastolic thresholds.¹ Nevertheless, these thresholds are not uniform and differ according to the clinical guidelines adopted globally. Among the most widely recognized are the Seventh Report of the Joint National Committee on Prevention, Detection, Evaluation, and Treatment of High Blood Pressure in Adults (JNC-7), the guidelines issued by the American College of Cardiology/American Heart Association (ACC/AHA), and those proposed by the International Society of Hypertension (ISH).^2,3,4^

For India, the latest National Family Health Survey categorizes Hypertension as mild, moderate or severely elevated blood pressure among adults 15 years and above.^5^ The globally accepted JNC-7 guidelines classify Hypertension in adults ≥18 years as pre-Hypertension, stage 1, and stage 2 Hypertension.^2^ The AHA and ISH guidelines also categorize HTN in various different ways.^3,4^ This difference in various classifications possibly results in ambiguity in interpretation of the data on prevalence of Hypertension and its associated risk factors.

Studies on hypertension and its risk factors in India are limited due to lack of regular and systematic data availability, with most evidence coming from periodic surveys such as the National Family Health Survey and selective programmatic reports.^6,7,8^ A comparison of these various guidelines, which we plan to do through this study, could result in identifying the gaps in the classification of Hypertension and would provide data, which is globally recognizable and of utility for the academia and policy makers.

## Methods

This study used secondary data from the fifth National Family Health Survey (NFHS-5, 2019– 20), coordinated by the International Institute for Population Sciences, Mumbai. NFHS-5 employed a stratified two-stage sampling design; detailed methodology is available in the survey report. Data were extracted from the women’s (n=6,29,294) and men’s (n=80,177) recode files.

Hypertension was defined as blood pressure ≥140/90 mmHg or self-report of being diagnosed with hypertension on at least two occasions, or current use of antihypertensive medication. To ensure comparability within the adult population, analyses were restricted to men aged 18–54 years and women aged 18–49 years. Pre-hypertensive (JNC-7: SBP 120–129 or DBP 80–89), elevated (AHA: SBP 120–129 and DBP <80), and high-normal (ISH: SBP 130–139 or DBP 85– 89) categories were excluded.

Data were cleaned in Microsoft Excel and analyzed using IBM SPSS version 29.0. Descriptive statistics summarized sociodemographic and clinical variables. Hypertension prevalence was estimated according to JNC-7, AHA, and ISH classifications and expressed as percentages with 95% confidence intervals (CIs), using the Clopper–Pearson exact method.

Multivariate logistic regression was performed to calculate adjusted odds ratios (AORs) with 95% CIs, considering hypertension as the dependent variable. Independent variables included sex, age, education, occupation, household size, wealth quintile, body mass index, waist–hip ratio, smoking, alcohol use, place of residence, and religion. Self-reported comorbidities (heart disease, diabetes, asthma) were also included. Analyses accounted for survey weights, and statistical significance was set at p<0.05

## Results

Based upon the NFHS-5 data, the prevalence of Hypertension in Indian adult males (≥ 18 years) according to the JNC-7 and ISH guidelines is 21.78% (95% CI: 21.49-22.07). The prevalence in females is 12.35% (95% CI: 12.27-12.43). According to the AHA guidelines, the prevalence in males is 62.55% (95%CI: 62.22-62.28) and in females it is 45.53 % (95%CI: 45.41-45.65). **(Table 1)**

**Table 1:** Prevalence of Hypertension in Indian adults according to different guidelines.

|  | **Males (18-54) (% ,95%CI)** | **Females (18-49) (% ,95%CI)** |
| --- | --- | --- |
| **JNC-7 (Stage 1 + stage2)** | 21.78 (21.49-22.07) | 12.35 (12.27-12.43) |
| **AHA (Stage 1+ stage 2+crisis)** | 62.55 (62.22-62.89) | 45.53 (45.41-45.65) |
| **ISH (Stage 1+ stage2+ crisis)** | 21.78 (21.49-22.07) | 12.35 (12.27-12.43) |
| **NFHS-5** | **Males (15-54)** | **Females (15-49) (% ,95%CI)** |
|  | 24% | 21.3% |

### Prevalence of HTN by background characteristics]

#### JNC-7 & ISH guidelines

According to the Seventh Report of the Joint National Committee on Prevention, Detection, Evaluation, and Treatment of High Blood Pressure in Adults (JNC-7), 46.4% of women and 25.9 % of men and are normotensive (SBP <120-139 and DBP <80).41.3% of women and 52.3% of men have prehypertension (SBP 120-139 or DBP 80-89).Among the females, 9.6% have stage 1(SBP 140-159 or DBP 90-99) and 2.8% have stage 2 HTN (SBP ≥160 or DBP ≥ 100). Among the males, 16.6% and 5.2% have stage 1 and stage 2 respectively.

According to the International Society of Hypertension (ISH) guidelines, 72.9% of women and 56.1 % of men and are normotensive (SBP <130 or DBP <85).14.8 % women and 22.2% men have high-normal BP (SBP 130-139 and/or DBP 85-89). Grade 1 hypertension (SBP 140-159 and/or DBP 90-99) has been observed in 9.6% females and in 16.6% males while grade 2 (SBP ≥160 and /or DBP ≥100) has been observed in 2.8% females and 5.2% males.

#### As per both JNC-7 and ISH

Among the age groups, highest prevalence has been observed in the 40+ age groups for both the genders (females: 24.08% and males: 33.45%) in both JNC-7 and ISH.

As per educational attainment, the highest prevalence of HTN has been observed in the illiterate (16.88%) and lowest in those who have been to higher secondary and above (9.27%).s Higher prevalence of hypertension has been observed in the religious minorities, both among females (Sikhs: 17.94%, Buddhists: 15.19%, Christians: 12.63%) and males (Sikhs: 32.39%, Buddhists: 25.79%, Christians: 23.26%).

Among the ethnic groups, higher prevalence of HTN has been observed in the scheduled tribes (13.66%) as compared to the schedules castes (11.75%) and other backward classes (11.79%).

According to the wealth index, the highest prevalence has been seen in those belonging to the richest class (13.48%) and lowest among the poorest (11.58%).Higher prevalence has been observed among the urbanites (13.53%) as compared to the rural population (11.98%).According to Body Mass Index, the obese (BMI>25) have higher prevalence (22.76%) than the non-obese (9.29%). Among the alcohol users, higher prevalence has been observed (25.14%) as compared to the non-users (11.73%). Smokers have higher prevalence of HTN (21.88%) than non-smokers (12.18%).

Among the co-morbid conditions, higher prevalence has been seen in diabetics (RBS>200) (39.18%) as compared to the non-diabetics (11.48%) and in those with Asthma (17.77%) as compared to those without the condition (12.28%).

### American Heart Association guidelines

As per the American Heart Association guidelines (2017 and 2025), 46.4% of women and 25.9 % of men and are normotensive (SBP <120-139 and DBP <80).8.1% of women and 11.5% of men have elevated BP (SBP 120-129 and DBP <80). Stage 1 hypertension (SBP 130-139 or SBP 80-89) has been observed in 33.2% females and 40.8% of the males). Stage 2 hypertension (SBP≥140 and DBP ≥90) is present in 11.9% of females and 21.1% males). Crisis stage (SBP >180 and/or DBP >120) has been observed in 0.4% of females and 0.7% males). Among the age groups, highest prevalence has been observed in the 40+ age groups for both the genders (females: 63.66% and males: 74.79%).

As per educational attainment, the highest prevalence of HTN has been observed in the illiterate (53.52%) and lowest in those who have been to higher secondary and above (40.50%). Higher prevalence of hypertension has been observed in the religious minorities, (Sikhs: 53.59%, Buddhists: 51.97%, Christians: 47.39%).

Among the ethnic groups, higher prevalence of HTN has been observed in the scheduled tribes (46.37%) as compared to the schedules castes (41.07%) and other backward classes (41.46%).

According to the wealth index, the highest prevalence has been seen in those belonging to the richest class (45.17%) and lowest among the poorest (41.79%).Higher prevalence has been observed among the urbanites (46.76%) as compared to the rural population (44.21%).According to Body Mass Index, the obese (BMI>25) have higher prevalence (62.89%) than the non-obese (39.48%). Among the alcohol users, higher prevalence has been observed (65.03%) as compared to the non-users (43.84%). Smokers have higher prevalence of HTN (62.40%) than non-smokers (44.52%).

Among the co-morbid conditions, higher prevalence has been seen in diabetics (RBS>200) (76.21%) as compared to the non-diabetics (44.10%) and in those with Asthma (51.05%) as compared to those without the condition (44.74%). **(Table 2)**

**Table 2:**
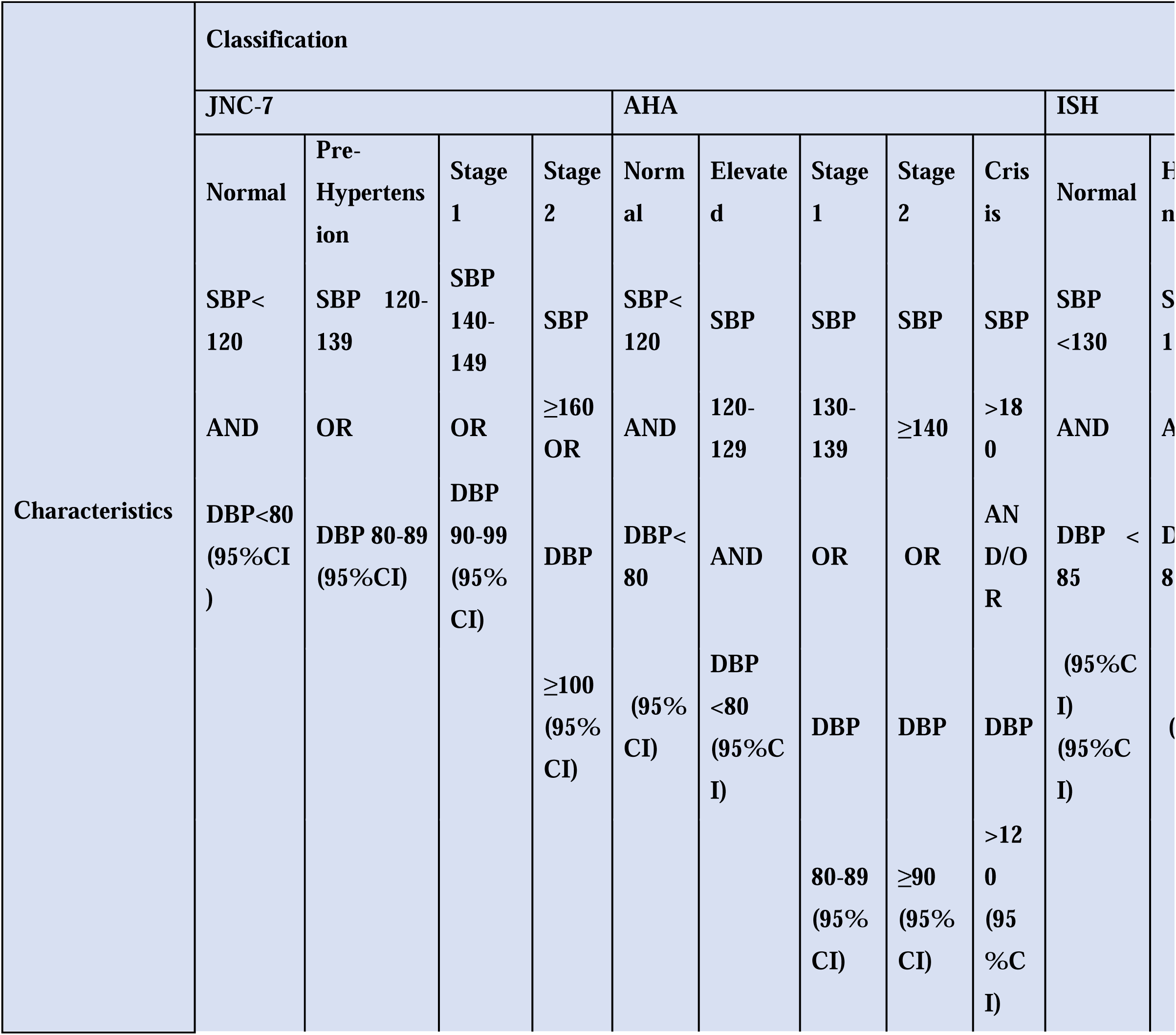
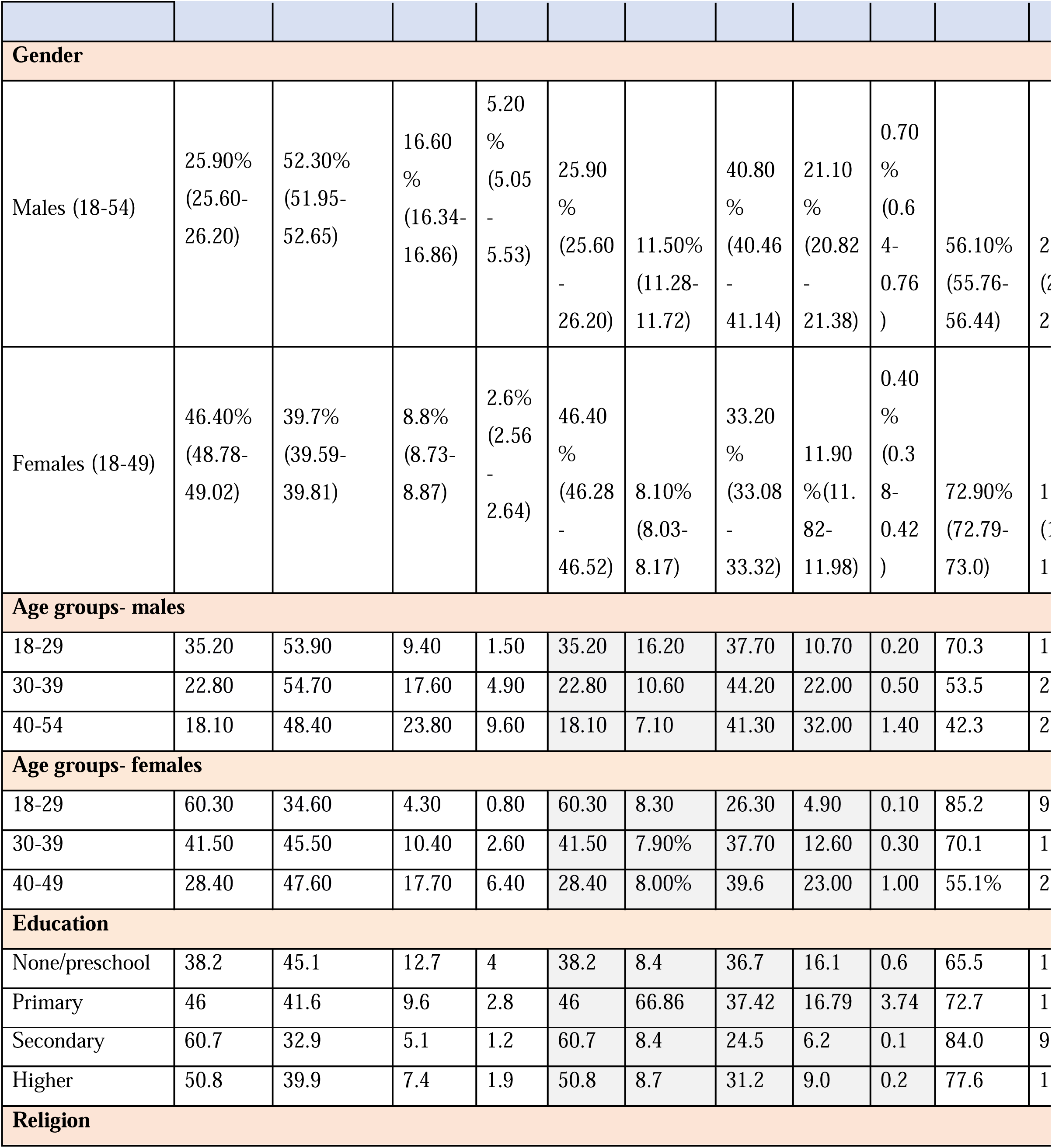
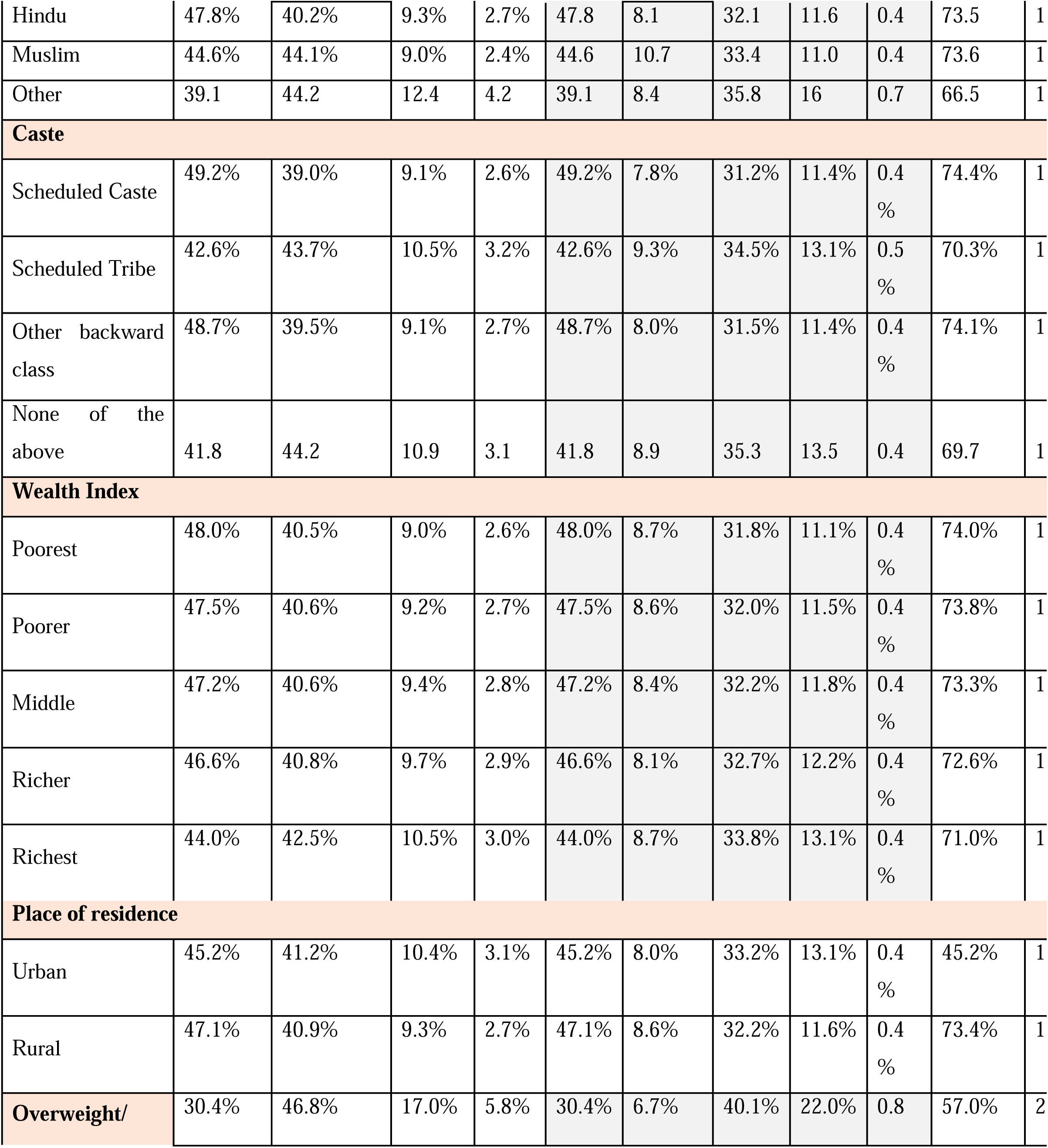
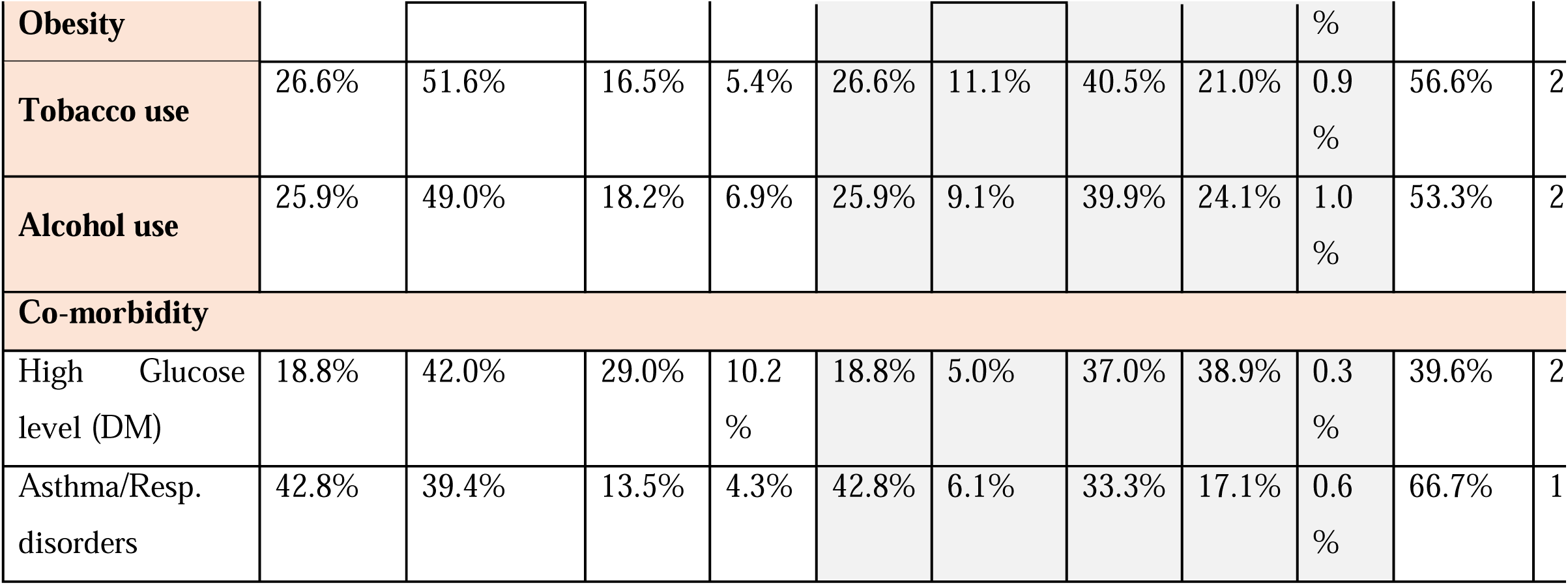
Prevalence of Hypertension by background characteristics in India, according to three separate guidelines.

The absolute differences in prevalence of Hypertension between JNC-7 and AHA classifications is 33.19% among females and 40.77% among males. **(Table 3)**

**Table 3:**
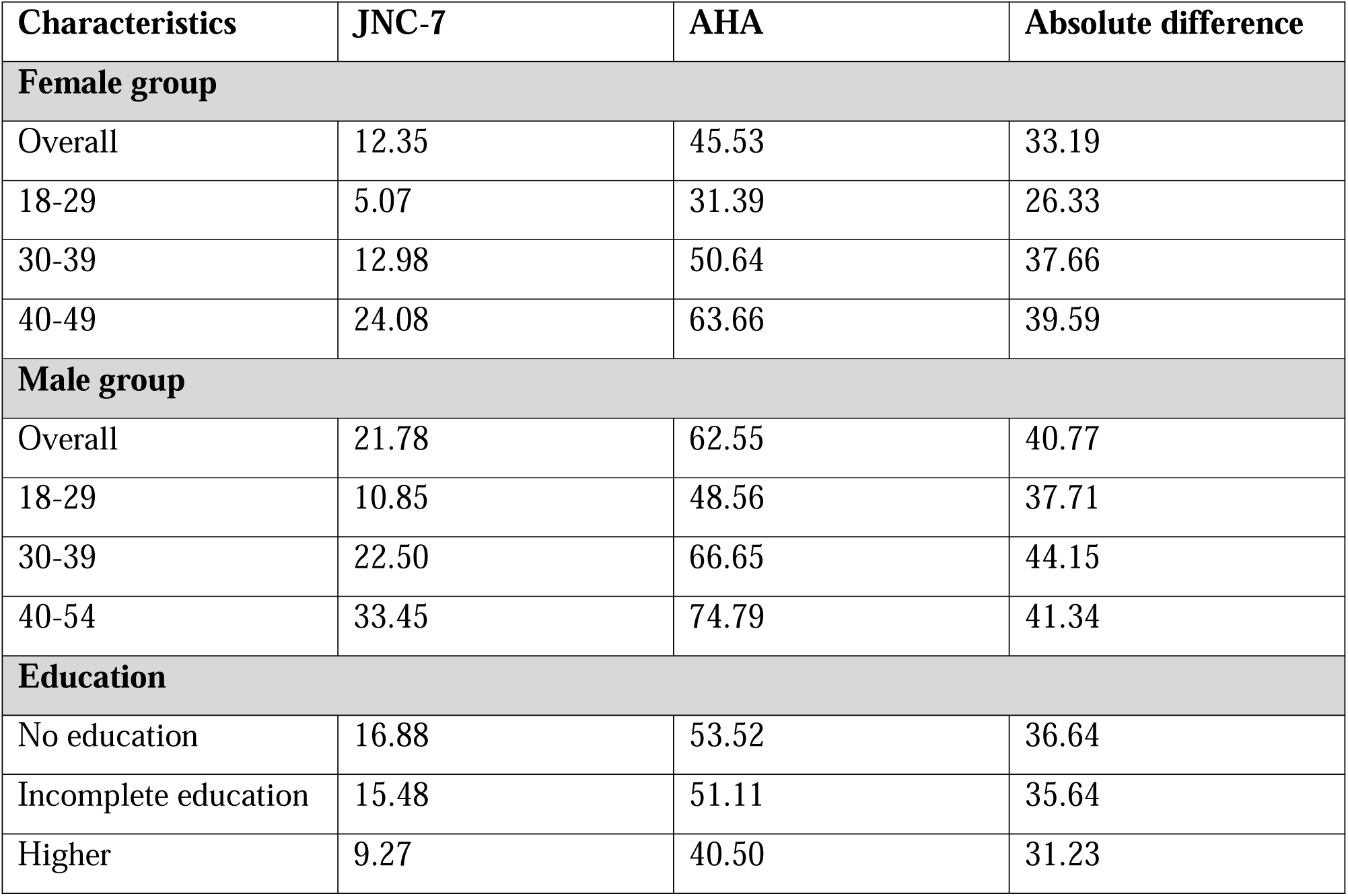
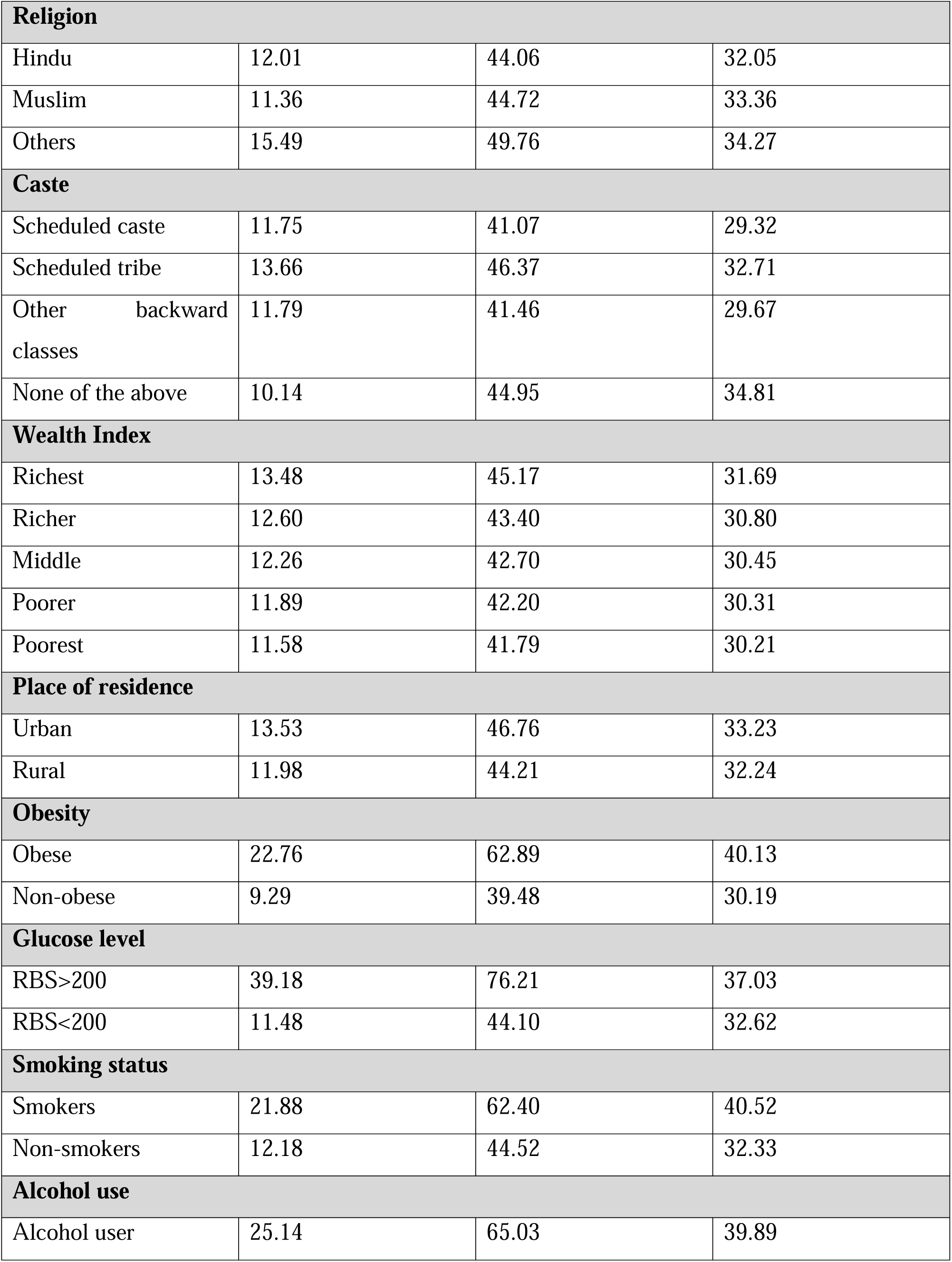
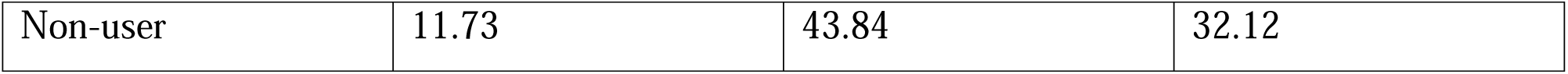
Table 2: Absolute changes in prevalence (%) of Hypertension by background characteristics.

As per logistic regression analysis among women, highest odds of having hypertension has been observed in the 40+ age groups across all the three guidelines.[JNC & ISH: AOR 3.99(3.90-4.02), AHA: AOR 3.02(2.98-3.07)]. Highest odds have been seen among the richest as per the AHA [AOR 1.03 (1.01-1.05) and the ISH guidelines [AOR 1.14 (1.08-1.21)]. Urbanites have been observed to have higher odds as compared to their rural counterparts as per the JNC-7 and ISH guidelines [AOR 1.24 (1.18-1.30). Smokers have higher odds as per JNC-7[AOR 1.46 (1.39-1.47), AHA [AOR 1.46 (1.41-1.52)] and ISH [AOR 1.52(1.45-1.59)]. Higher odds of having hypertension have also been observed among the alcohol users as per JNC-7 [AOR 1.46(1.39-1.47)], AHA [AOR 1.46(1.41-1.52)] and ISH [AOR 1.52(1.45-1.59)]. Diabetics also have higher odds as per JNC-7 [AOR 1.65(1.60-1.71), AHA [AOR 1.65(1.60-1.70) and ISH [AOR 2.20 (2.12-2.27)]. **(Table 4)**

**Table 4:** Results of Bivariate logistic regression analysis investigating risk factors for Hypertension (JNC-7 guideline) among men. NFHS-5 (2019-21)

|  | **JNC-7** | **AHA** | **ISH** |
| --- | --- | --- | --- |
| **Background Characteristics** | **AOR with 95% CI** | **AOR with 95% CI** | **AOR with 95% CI** |
| **Age Group** |  |  |  |
| 18-29® |  |  |  |
| 30-39 | 1.96*** (1.86, 2.06) | 1.85*** (1.78-1.92) | 1.96*** (1.86, 2.06) |
| 40-54 | 3.36*** (3.20, 3.53) | 2.73*** (2.63-2.84) | 3.36*** (3.20, 3.53) |
| **Obesity** |  |  |  |
| No ® |  |  |  |
| Yes | 2.02*** (1.94, 2.10) | 2.16*** (2.07-2.25) | 2.02*** (1.94, 2.10) |
| **Education** |  |  |  |
| No Formal Education® |  |  |  |
| Primary | 1.06 (0.99, 1.14) | 1.01 (0.95-1.07) | 1.06 (0.99, 1.14) |
| Secondary | 1.09*** (1.02, 1.15) | 1.05* (1.00-1.11) | 1.09*** (1.02, 1.15) |
| Higher | 1.06 (0.98, 1.13) | 1.11*** (1.04-1.18) | 1.06 (0.98, 1.13) |
| **Wealth Index** |  |  |  |
| Poorest® |  |  |  |
| Poorer | 1.01 (0.96, 1.08) | 1.05* (1.00-1.10) | 1.01 (0.96, 1.08) |
| Middle | 1.08** (1.01, 1.14) | 1.04 (0.99-1.09) | 1.08** (1.01, 1.14) |
| Richer | 1.09*** (1.02, 1.16) | 1.09*** (1.04-1.15) | 1.09*** (1.02, 1.16) |
| Richest | 1.19*** (1.11, 1.27) | 1.21*** (1.15-1.28) | 1.19*** (1.11, 1.27) |
| **Place of Residence** |  |  |  |
| Rural® |  |  |  |
| Urban | 1.14*** (1.09, 1.19) | 1.16*** (1.12-1.20) | 1.14*** (1.09, 1.19) |
| **Tobacco Use** |  |  |  |
| No® |  |  |  |
| Yes | 0.99*(0.95 , 1.04) | 1.04 (0.99, 1.08) | 0.99*(0.95 , 1.04) |
| **Alcohol Use** |  |  |  |
| No® |  |  |  |
| Yes | 1.43*** (1.38 , 1.49) | 1.39*** (1.34-1.44) | 1.43*** (1.38 , 1.49) |
| **Diabetes** |  |  |  |
| No® |  |  |  |
| Yes | 1.84*** (1.71 , 1.98) | 1.43*** (1.31-1.55) | 1.84*** (1.71 , 1.98) |
| AOR: Adjusted odd ratio, ®<br>Reference category;<br>*p<0.05; **p<0.005;<br>***p<0.001 |  |  |  |
| **AOR: Adjusted odd ratio; ® Reference category; \*p<0.05; \*\*p<0.005; \*\*\*p<0.001** |  |  |  |

As per logistic regression analysis among men, highest odds of having hypertension have been observed among the 40-54 age group across all the three guidelines [JNC & ISH: AOR 3.36(3.20-3.53), AHA: AOR 2.73(2.63-2.84)]. The obese have also been observed to have the higher odds than non-obese as per JNC-7 & ISH [AOR 2.02(1.94-2.10)] and AHA [AOR 2.16(2.07-2.25)]. Highest odds have been observed among those educated up to the secondary level as per JNC-7 and ISH [AOR 1.09(1.02-1.15)] and those educated above higher secondary as per the AHA [AOR 1.11(1.04-1.18)].The wealthiest have shown to have the highest odds of hypertension as per all the three guidelines [JNC & ISH: AOR 1.19(1.11-1.27 and AHA: AOR 1.21(1.15-1.28)]. The urbanites have also shown to have the higher odds as compared to their rural counterparts as per all three guidelines [JNC & ISH: AOR 1.14(1.09-1.19 and AHA: AOR 1.16(1.12-1.20)]. Alcohol users and diabetics have also shown to have the higher odds across all three guidelines [Alcohol users: JNC & ISH: AOR 1.43(1.38-1.49 and AHA: AOR 1.39(1.34-1.44)] [Diabetics: JNC & ISH: AOR 1.84(1.71-1.98 and AHA: AOR 1.43(1.31-1.55)]. Smokers have shown to have the higher odds than non-smokers as per the AHA guidelines [AOR 1.04(0.99-1.08), however, these results are not statistically significant. **(Table 5)**

**Table 5:** Results of Bivariate logistic regression analysis investigating risk factors for Hypertension (JNC-7 guideline) among women. NFHS-5 (2019-21)

|  | **JNC-7** | **AHA** | **ISH** |
| --- | --- | --- | --- |
| **Background Characteristics** | aOR (95% CI) |  |  |
| **Age Group** |  |  |  |
| 18-29® |  |  |  |
| 30-39 | 2.05*** (2.01,2.10) | 1.89*** (1.87,1.92) | 2.05*** (2.01,2.10) |
| 40-49 | 3.99*** (3.90,4.02) | 3.02*** (2.98,3.07) | 3.99*** (3.90,4.02) |
| **Obesity** |  |  |  |
| No ® |  |  |  |
| Yes | 2.09*** (2.05 , 2.12) | 1.96*** (1.93,1.99) | 2.09*** (2.05 , 2.12) |
| **Education** |  |  |  |
| No Formal Education® |  |  |  |
| Primary | 1.01 (0.97 , 1.03) | 1.01(0.99,1.03) | 1.01 (0.97 , 1.03) |
| Secondary | 0.89*** (0.87 , 0.91) | 0.88*** (0.87,0.89) | 0.89*** (0.87 , 0.91) |
| Higher | 0.72*** (0.7 , 0.74) | 0.77*** (0.76,0.79) | 0.72*** (0.7 , 0.74) |
| **Wealth Index** |  |  |  |
| Poorest® |  |  |  |
| Poorer | 1.02 (0.99 , 1.05) | 1.00(0.99,1.02) | 1.01(0.98,1.03) |
| Middle | 0.98(1.01 , 1.07) | 0.99(0.98,1.01) | 0.98(0.96,1.01) |
| Richer | 0.97** (1.01 , 1.07) | 1.00(0.98,1.02) | .97(.94,1.00) |
| Richest | 1.01 (0.98 , 1.04) | 1.03*** (1.01,1.05) | 1.14** (1.08,1.21 ) |
| **Place of Residence** |  |  |  |
| Rural® |  |  |  |
| Urban | 1.02** (1.01 , 1.04) | 1.00(0.99,1.02) | 1.02** (1.00,1.04 ) |
| **Tobacco Use** |  |  |  |
| No® |  |  |  |
| Yes | 1.24*** (1.18 , 1.30) | 1.19*** (1.15,1.23) | 1.24*** (1.18 , 1.30) |
| **Alcohol Use** |  |  |  |
| No® |  |  |  |
| Yes | 1.46*** (1.46 , 1.41) | 1.46*** (1.41,1.52) | 1.52*** (1.45,1.59) |
| **Diabetes** |  |  |  |
| No ® |  |  |  |
| Yes | 1.65***(1.60 , 1.71) | 1.65***(1.60,1.70) | 2.20***(2.12,2.27) |
| AOR: Adjusted odd ratio; ® Reference category; *p<0.05; **p<0.005; ***p<0.001 |  |  |  |

## Discussion

Our study performed an innovative and thorough comparison of hypertension prevalence within the Indian context by utilizing three established diagnostic criteria: the International Society of Hypertension (ISH), the Joint National Committee (JNC), and the American Heart Association (AHA) guidelines. Although each of these scales is founded on evidence-based thresholds, they exhibit both nuanced and significant differences that can substantially impact diagnostic and therapeutic choices. These differences are not just theoretical; they have practical effects on how patients are treated. For instance, a person labelled as “hypertensive” according to one guideline may concurrently be designated as “high normal” or “pre-hypertensive” according to another, despite having identical blood pressure measurements. Our study used a large dataset from NFHS-5 that was representative of the whole country to show how much this diagnostic variability exists. This shows how different clinical paths and treatment choices can be made just by choosing a scale.

### Prevalence of Hypertension

The NFHS-5 reports the overall prevalence of hypertension in adults (15 years and above) as 21.3% in females and 24% in males. While applying the JNC-7 and ISH cut-offs, hypertension in adults (≥ 18 years) the prevalence has been calculated to be 12.35% in females and 21.78% in males. According to the AHA cut-offs, the prevalence comes out to be 45.53% in males and 62.55% in males. Several authors have highlighted that discrepancies in the classification of hypertension across guidelines, such as JNC-7, AHA, and ISH, may create challenges for both clinical decision-making and research comparability.^9,10,11,12^

## Background characteristics

### Socio-demographic determinants

Sociodemographic factors strongly influence the distribution of hypertension in India. Age remains the most important non-modifiable determinant, with prevalence rising sharply after 40 years in both men and women.^13^ In our study too, highest odds of having hypertension have been observed among the 40+ age group in both the sexes according to all three guidelines. Men generally exhibit higher prevalence than women, though the gap narrows with advancing age.^14^ In the present study too, prevalence in men has been observed to be 9.4% more than women as per JNC-7 and ISH guidelines and 17.02% more in men than women as per AHA guidelines. Educational attainment appears protective, with systematic reviews showing significantly lower odds of hypertension among the better educated, likely reflecting enhanced awareness and healthier behaviors.^15^ Our study also highlights the significantly lower odds of hypertension among the better educated. Socioeconomic gradients are also evident, as national surveys such as NFHS-4 and NNMS report higher odds of hypertension among individuals in the richest quintiles compared with the poorest.^5,16^ This study also highlights the highest odds of having hypertension among the richest class. Urban residence consistently confers greater risk relative to rural settings, underscoring the influence of urbanization, lifestyle changes, and environmental exposures.^15^ Our study also highlights that urbanites have the higher odds of having hypertension as compared to their rural counterparts. Collectively, these sociodemographic associations highlight the dual role of structural and individual-level factors in shaping the hypertension burden.

### Behavioral/metabolic determinants

Behavioral and metabolic risk factors play a pivotal role in shaping the hypertension burden in India. Excess body weight is a critical risk factor, with Indian adults having BMI ≥25 kg/m² showing two-to three-fold higher odds of hypertension.^17^ Our study also highlights about two-fold higher odds in those with a BMI>25. Hypertension also clusters strongly with diabetes, with systematic reviews indicating a two-to three-fold greater prevalence among diabetics compared with non-diabetics, a pattern further corroborated by national survey data.^18^ Our study also re-affirms this association across all guidelines. Alcohol consumption has been linked with greater hypertension prevalence, emphasizing its role as a modifiable determinant.^19^ In our study too, higher odds of having hypertension have been observed in alcohol users across all guidelines. Tobacco use has been consistently associated with elevated blood pressure, with pooled evidence showing higher prevalence among smokers compared to nonsmokers, and national surveys such as NNMS (2017–18) confirming significantly higher odds of hypertension among ever-smokers.^20,21^ Higher odds of hypertension have been observed in our study too among female smokers. However, this association is statistically non-significant in case of males, The lower odds ratio observed among males may be attributed to the smaller subgroup sample size. This may also be due to ‘smoker’s paradox’ as cited in a recent study.^22^

Collectively, these findings underscore the urgent need for integrated NCD control strategies in India, targeting shared behavioral and metabolic risk factors.

### Need for a uniform classification system

Existing hypertension prevalence data through surveys like NFHS and globally accepted guidelines including JNC-7, AHA 2017, and ISH 2020, differ in their classification thresholds and staging criteria. These variations result in different prevalence estimates and may influence treatment decisions, thereby creating uncertainty in clinical practice. Several authors have emphasized that the adoption of a uniform classification system would be beneficial, as it would allow for consistent treatment protocols, facilitate international comparability of research findings, and strengthen public health policies.^23,24,25,26^ In this context, our study compared the prevalence of hypertension across these commonly used guidelines to highlight the magnitude of variation and underscore the need for harmonization in classification systems.

Our findings reinforce the need for a uniform hypertension classification framework, which would not only improve consistency in clinical decision-making but also aid policymakers in formulating standard treatment guidelines and comparable surveillance strategies.

## Strengths and Limitations

One of the major strengths of our study lies in the use of NFHS-5, a nationally representative and large-scale dataset that lends strong external validity to our findings. To the best of our knowledge, this is the first analysis in the Indian context that directly compares the ISH, JNC, and AHA hypertension criteria, thereby filling an important gap in the literature. By systematically examining subcategories such as normal, high normal, pre-hypertensive, and stage 1 or stage 2 hypertension, our study was able to highlight the subtle as well as substantial variations across these guidelines. Importantly, the implications of our findings go well beyond differences in prevalence estimates—they extend to clinical practice, health policy framing, and, most crucially, to patient-level outcomes in the Indian healthcare setting.

Our study does have some flaws, though. NFHS-5 relies on cross-sectional blood pressure measurements, which may inadequately capture intra-individual variability or the impact of ‘white-coat’ effects. Moreover, the lack of longitudinal follow-up inhibits our capacity to determine whether the observed diagnostic variations result in differences in cardiovascular outcomes. Another limitation is that we could not account for treatment history in detail, which may have affected classification and prevalence estimates across the various scales.

## Conclusions

Our findings demonstrate substantial variation in hypertension prevalence estimates depending on the classification criteria applied, reflecting the inconsistencies across NFHS, JNC-7, AHA 2017/2025, and ISH 2020 guidelines. This variability has implications for clinical practice, research comparability, and public health policy. Adoption of a uniform hypertension classification framework would strengthen the reliability of prevalence estimates, ensure equitable treatment decisions, and enable policymakers to design standardized, evidence-based strategies. Importantly, such harmonization is essential to accelerate progress towards the Sustainable Development Goal (SDG) target of reducing the prevalence of hypertension and mitigating its associated health burden by 2030.

## Recommendations

1. **Adopt a Uniform Hypertension Classification Framework**: Policymakers and professional societies should work towards consensus on thresholds and staging criteria that balance clinical applicability with global comparability.
2. **Integrate Standardized Guidelines into National Programs**: India’s hypertension control initiatives (such as NPCDCS and IHIP) should incorporate a uniform definition to ensure consistency in screening, diagnosis, and management.
3. **Enhance Surveillance Systems**: National surveys like NFHS should align their definitions with globally agreed cut-offs, enabling more reliable tracking of prevalence trends and progress toward SDG targets.
4. **Promote Capacity Building and Awareness**: Training of healthcare providers and community workers in the standardized classification will minimize variability in diagnosis and treatment.
5. **Facilitate International Comparability**: Harmonized definitions will make Indian data directly comparable with global evidence, aiding in research collaboration and policy formulation.

## Data Availability

All data produced in the present study are available upon reasonable request to the authors

